# Assessment of COVID-19 Effect on the Health of Families in South-West, Nigeria

**DOI:** 10.1101/2022.08.10.22278638

**Authors:** O Olorunfemi, O.O Oluwagbemi, O.E Atekoja, A.O Olajide, O.O Olu-Abiodun, J.A Adebiyi, J.O Sodimu, T.A Leslie, E.A Ojo, M.O Akpa

**Author notes:** **Correspondence: Olorunfemi**.O. @, 08034694675.

## Abstract

**Aim:** This study was to assess the effect of the coronavirus disease and its associated lockdown on the physical, spiritual, emotional, and socio-economic health of families living in South-West, Nigeria.

**Background:** The outbreak of the COVID-19 pandemic create a universal health crisis that has a major effect on our day–to-day activities and these global concerns have shifted from the diseases to the physical, emotional, spiritual, and socioeconomic effects of the situation on the people.

**Method:** This is a descriptive study with five hundred and thirty-six (536) respondents; a convenient sampling technique was used to select samples through online Google form.

**Result:** The majority of the respondent’s ages ranged between 20 and 30years (53.0%). COVID 19 pandemic affected 17.2% of the respondents’ physical health. The lockdown improved bonding among family members (74.6%), also the lockdown favourable affected 56.0% of the respondents’ emotional health. The spirituality of the respondents was negatively affected (79.9%) by the lockdown, family expenses were increased (82.6%), there was an increased in the prices of goods (92.9%), and in general COVID 19 pandemic unfavorably affected (77.2%) the respondents’ socio-economic health.

**Conclusion:** This study reveals that COVID-19 and the lockdown produced an adverse effect on the physical, emotional, spiritual, and socio-economic wellbeing of the families in southwest Nigeria.

**Implications for nursing and health policy:** nurses working in COVID-19 unit need to give total care to the affected patient; therefore, they have obligation to include physical, emotional, spiritual and mental intervention in their care. The government needs to plan and strategize properly in the method for distribution of the palliative’s, and if possible identify the vulnerable and less privileged in each state for easy distribution.

## Introduction

The coronavirus diseases otherwise known as COVID-19, started on the 31^st^ of December, 2019 in Wuhan, China, and spread to 212 countries of the world (Johns Hopkins University, 2020). The spread of the disease continues and on 12^th^ of March, 2020, the World Health Organization (WHO) declared the disease as a pandemic (WHO, 2020) requiring countries to adopt appropriate measures. As at the 7^th^ of May, 2020, about 3.6 million confirmed cases of COVID-19, including 251,446 deaths was reported (WHO, 2020b).

On 27^th^ of February 2020, Nigeria reported the first case of coronavirus diseases, when one native of Italy visited Lagos state was found to be positive for COVID-19 (Agusi et al., 2020). The second incident of the disease was reported in Ogun State, on the 9^th^ of March 2020, this time; it was the citizen of Nigeria who had direct contact with the Italian man. The Nigeria government gives a public broadcast on 28 of January 2020 to assure the people of her country of the mechanism and systems put in place to contain and prevent the spread of diseases (Saha & Dutta, 2020).

In Nigeria study shows that the impact of coronavirus disease cases varies from one state to another, for example in Kogi the incidence is 0.09 per 100,000 compared to 83.7 per 100,000 in Lagos state (Hassan et al., 2020). However, about 70% of the cases of COVID-19 were majorly in seven States of the country: Lagos, Abuja, Plateau, Kaduna, Oyo, Kano, and Edo state. The mortality rate of the people affected by the pandemic also differs as one moves from one state to another, with Lagos, Abuja, and Plateau occupying the first position in the rating scale, as at the 7th of February, 2021.

Nigeria’s Federal Ministry of Health is saddle with the responsibility of put up measures aimed at controlling and containing the outbreak of any pandemic in Nigeria. These measures are to be synchronized and implemented by the Nigeria Centre for Disease Control (NCDC) an agency responsible for the investigation, monitoring, detection, prevention, and control of infectious diseases. Other measures such as the closure of Nigeria borders were implemented to contain the spread of the pandemic. However, in spite of these measures, the coronavirus disease continues to spread in the country with a total 139,242 confirmed cases, 112, 557 recovered and 1,647 deaths as of February 07, 2021. (NCDC Report). In lieu of this, through the Quarantine Act 1926; an Infectious Diseases Law, Nigeria government declared a state of emergency against the disease. The Act empowers the president to introduce measures that will prevent the introduction, spreading and transmission of the diseases from Nigeria to other neighboring countries. The President in pursuance of the Act the federal government declared the disease as an infectious and dangerous disease and made lockdown orders in three states of Nigeria; Abuja (FCT), Ogun, and Lagos states, and this was later extended to other states by the respective State Governors.

The disruption caused by the pandemic is overwhelming, and tens of millions of families are at risk of falling into poverty, and a high percentage of family members may lose their job. The disease creates global health challenges that have had a deep impact on families, which is the smallest unit of any country (Kolodko, 2020). Therefore it is imperative to assess the effect of the pandemic on the physical, mental, and socio-economic of families, especially for low-income and vulnerable groups.

### Aims/objective of the study

This study aim to assess the effect of COVID-19 on the physical, emotional, spiritual, and socio-economic health of families living in South-West, Nigeria

## Methods

This study was carried out using descriptive research design because it helps to describe specific phenomenon or to find relationship among variables discussed. This research was conducted in States in the South-west, Nigeria. South-West Nigeria is one of the geopolitical zones of Nigeria consisting of Oyo, Osun, Ondo, Ekiti, Lagos, and Ogun States. It is majorly a Yoruba speaking area. The target population of the study was families living in South-West, Nigeria. Convenient sampling technique was used to select samples. The Google form instruments were sent to families platforms such as WhatsApp, Facebook, telegram, and Twitter in the South-West for them to responds to the questions in the instrument.

### Data Collection

A self-structured instrument was used in the data collection, and this was divided into five sections (A, B, C, D, & E). It consists of thirty seven questions in all. Section A asks questions on Socio-Demographic characteristics of respondents and it has ten questions. Section B asks questions on the effects of Covid-19 on physical health of families in South-West, Nigeria and it consist of eight questions. Section C: asks questions on the effects of the diseases on emotional health of families in South-West, Nigeria and it consist of six questions. Section D asks questions on effects of Covid-19 on spiritual health of families in South-West, Nigeria and it consist of four questions. Section E asks questions on effects of Covid-19 on Socio-economic health of families in South-West, Nigeria, and it consist of seven questions. The instrument was shown to an expert in the field and statistician for content validation. Then a pilot study was carried out among families in two of the states in South-East using 50 participants through the use of Google forms. Cronbach’^s^ alpha test was carried out to test the internal consistency or reliability of the instrument, and correlational coefficient value of 0.8 was obtained, showing a high level of reliability of the instrument.

### Data Collection Method

A permission to collect data was collect from Babcock university ethical committee. The study’s purpose was clearly stated on the instrument through the google form. The researchers distribute the instrument to the families in the South west by sending the google form to different platforms of families such as WhatsApp, Facebook, telegram, and Twitter in the South-West. Data collection was done for two weeks after which the data was collated and analyzed.

### Data Analysis

The Data gotten from this study were analyzed using Statistical Package for the Social Sciences (SPSS); a statistical software version 21.00 (IBM corp released 2012 Armonk, NY: IBM Corp). Descriptive statistics of frequencies and percentages was used to answer the research questions. The researchers decide to use just descriptive statistical analysis to focus on the study aim and objectives, which is to determine the effect of corona virus disease on the physical, emotional, spiritual and socio-economic health of families and they don’t intent to look at relationship between the identified variables, that will required other inferential statistics.

### Ethical Approval

Ethical clearance was taking from Babcock University (GEDS-901) and Consent was sought from the respondents that wish to fill the instrument. The participants were informed of their right to refuse participation in the study without it having a negative impact on them. Only participants that consent and that want to participate voluntarily should only filled the form and to sign informed consent for voluntary participation in the research. Confidentiality was maintained as elements for research was instructed not to write their names to prevent identifications. They were assured that whatever information given will be treated privately, hence and they were advised and encourage answering the questions sincerely.

## Results

The socio-demographic characteristic of the participants is show in table 1. This study found that the coronavirus pandemic affect the physical and emotional health of families (table 2) Moreover, it also shows that the pandemic has major effect on the spiritual and socio-economic health of families (table 3). The novel disease is impacting every family at various level and aspects, some families are more severe than others. For a number of families, it could mean making changes in their day to day activities, as a result of the financial difficulty associated with the pandemic. For other families, it results into emotional and psychological problem like panic anxiety or general fear in the home. In most cases, the pandemic produce psychological distress and concerns for parents especially during the period of lockdown, due to unstable financial circumstances, and school closures.

**Table 1:** Demographic Characteristics of the Respondents.

| Variable | Category | Frequency (n) | Percentage (%) |
| --- | --- | --- | --- |
| Age group (yrs.) | 20 – 30 | 284 | 53.0 |
|  | 31 – 40 | 129 | 24.1 |
|  | 41 – 50 | 80 | 14.9 |
|  | 51 and Above | 43 | 8.0 |
| Gender | Female | 390 | 72.8 |
| Sex ratio = 0.37 | Male | 146 | 27.2 |
| Tribe | Yoruba | 425 | 79.3 |
|  | Igbo | 55 | 10.3 |
|  | Hausa | 3 | 0.6 |
|  | Others | 53 | 9.8 |
| Family Type | Extended | 53 | 9.9 |
|  | Nuclear | 429 | 80.0 |
|  | Single parent | 54 | 10.1 |
| Family size | 1 – 2 | 40 | 7.5 |
|  | 3 – 4 | 193 | 36.0 |
|  | 5 – 6 | 201 | 37.5 |
|  | 7 – 8 | 66 | 12.3 |
|  | Above 8 | 36 | 6.7 |
| Educational status | No formal | 4 | 0.7 |
|  | Primary | 7 | 1.3 |
|  | Secondary | 14 | 2.6 |
|  | Tertiary | 511 | 95.3 |
| Employment status | Government employee | 128 | 23.9 |
|  | Private employee | 118 | 22.0 |
|  | Self employed | 137 | 25.6 |
|  | Unemployed | 153 | 28.5 |
| Place of Residence | No response | 3 | 0.6 |
|  | Ekiti | 76 | 14.2 |
|  | Lagos | 155 | 28.9 |
|  | Ogun | 157 | 29.3 |
|  | Ondo | 48 | 9.0 |
|  | Osun | 25 | 4.7 |
|  | Oyo | 72 | 13.4 |
| Travel history before lockdown | No | 345 | 64.4 |
|  | Yes | 191 | 35.6 |
| COVID 19 status | I don't know | 125 | 23.3 |
|  | Negative | 408 | 76.1 |
|  | Positive | 3 | 0.6 |

**Table 2:**
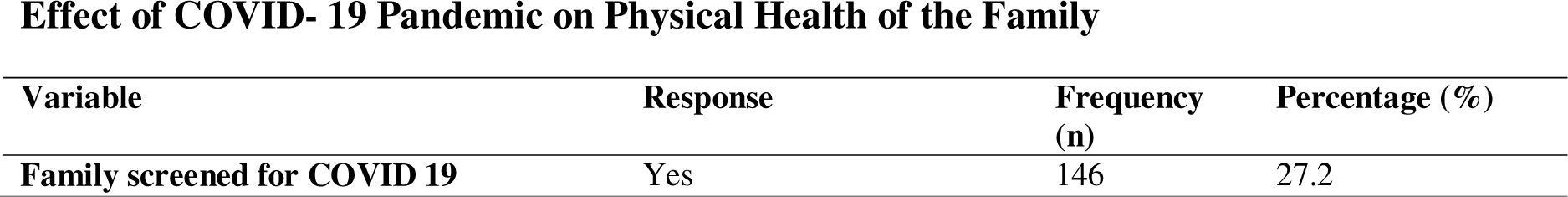
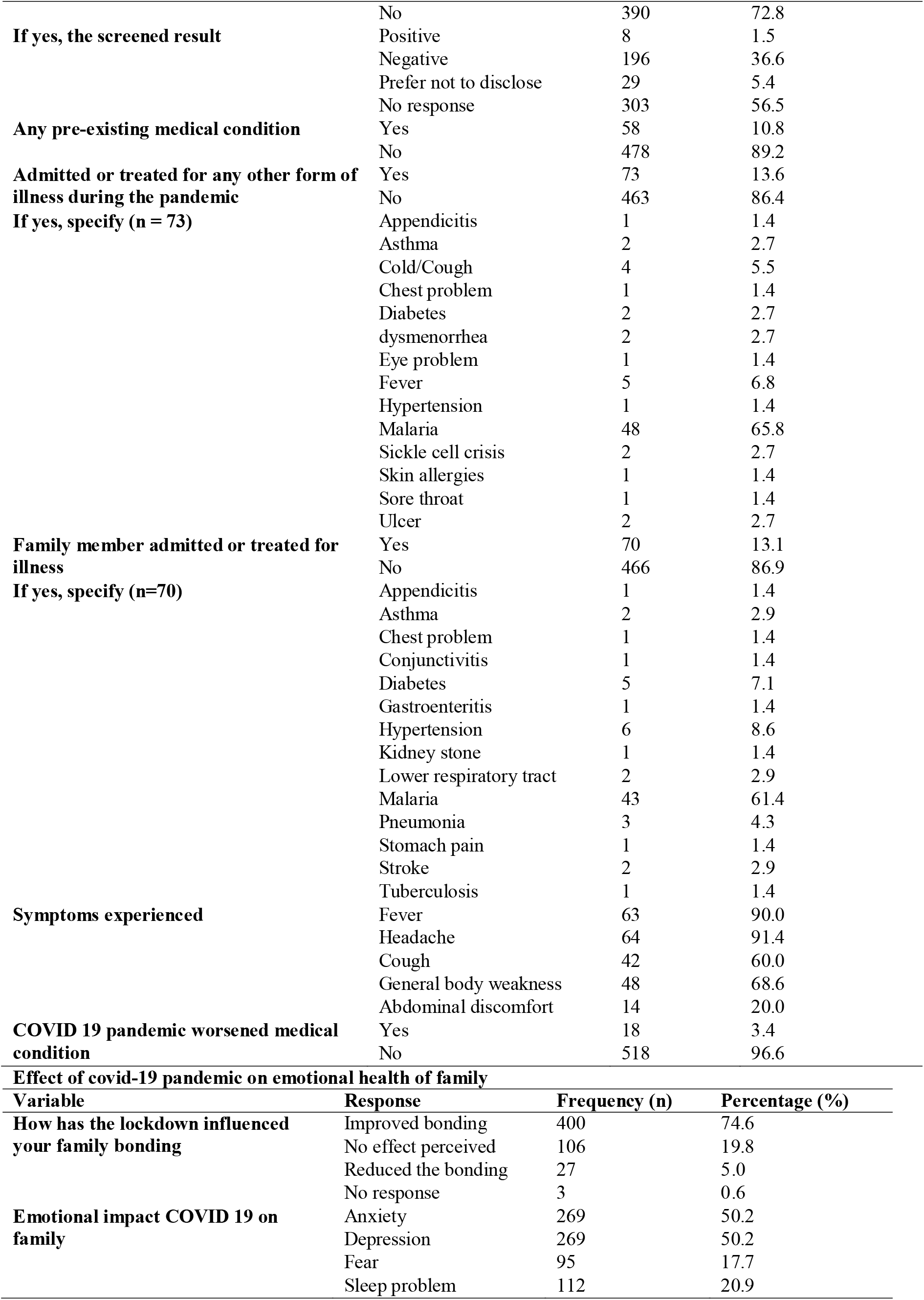
EFFECT OF COVID 19 PANDEMIC ON PHYSICAL AND EMOTIONAL HEALTH OF THE FAMILY.

Effect of COVID- 19 Pandemic on Physical Health of the Family
| Variable | Response | Frequency (n) | Percentage (%) |
| --- | --- | --- | --- |
| Family screened for COVID 19 | Yes | 146 | 27.2 |
|  | No | 390 | 72.8 |
| **If yes, the screened result** | Positive | 8 | 1.5 |
|  | Negative | 196 | 36.6 |
|  | Prefer not to disclose | 29 | 5.4 |
|  | No response | 303 | 56.5 |
| **Any pre-existing medical condition** | Yes | 58 | 10.8 |
|  | No | 478 | 89.2 |
| **Admitted or treated for any other form of illness during the pandemic** | Yes | 73 | 13.6 |
|  | No | 463 | 86.4 |
| **If yes, specify (n = 73)** | Appendicitis | 1 | 1.4 |
|  | Asthma | 2 | 2.7 |
|  | Cold/Cough | 4 | 5.5 |
|  | Chest problem | 1 | 1.4 |
|  | Diabetes | 2 | 2.7 |
|  | dysmenorrhea | 2 | 2.7 |
|  | Eye problem | 1 | 1.4 |
|  | Fever | 5 | 6.8 |
|  | Hypertension | 1 | 1.4 |
|  | Malaria | 48 | 65.8 |
|  | Sickle cell crisis | 2 | 2.7 |
|  | Skin allergies | 1 | 1.4 |
|  | Sore throat | 1 | 1.4 |
|  | Ulcer | 2 | 2.7 |
| **Family member admitted or treated for illness** | Yes | 70 | 13.1 |
|  | No | 466 | 86.9 |
| **If yes, specify (n=70)** | Appendicitis | 1 | 1.4 |
|  | Asthma | 2 | 2.9 |
|  | Chest problem | 1 | 1.4 |
|  | Conjunctivitis | 1 | 1.4 |
|  | Diabetes | 5 | 7.1 |
|  | Gastroenteritis | 1 | 1.4 |
|  | Hypertension | 6 | 8.6 |
|  | Kidney stone | 1 | 1.4 |
|  | Lower respiratory tract | 2 | 2.9 |
|  | Malaria | 43 | 61.4 |
|  | Pneumonia | 3 | 4.3 |
|  | Stomach pain | 1 | 1.4 |
|  | Stroke | 2 | 2.9 |
|  | Tuberculosis | 1 | 1.4 |
| **Symptoms experienced** | Fever | 63 | 90.0 |
|  | Headache | 64 | 91.4 |
|  | Cough | 42 | 60.0 |
|  | General body weakness | 48 | 68.6 |
|  | Abdominal discomfort | 14 | 20.0 |
| **COVID 19 pandemic worsened medical condition** | Yes | 18 | 3.4 |
|  | No | 518 | 96.6 |
| **Effect of covid-19 pandemic on emotional health of family** |  |  |  |
| **Variable** | **Response** | **Frequency (n)** | **Percentage (%)** |
| **How has the lockdown influenced your family bonding** | Improved bonding | 400 | 74.6 |
|  | No effect perceived | 106 | 19.8 |
|  | Reduced the bonding | 27 | 5.0 |
|  | No response | 3 | 0.6 |
| **Emotional impact COVID 19 on family** | Anxiety | 269 | 50.2 |
|  | Depression | 269 | 50.2 |
|  | Fear | 95 | 17.7 |
|  | Sleep problem | 112 | 20.9 |

**Table 3:**
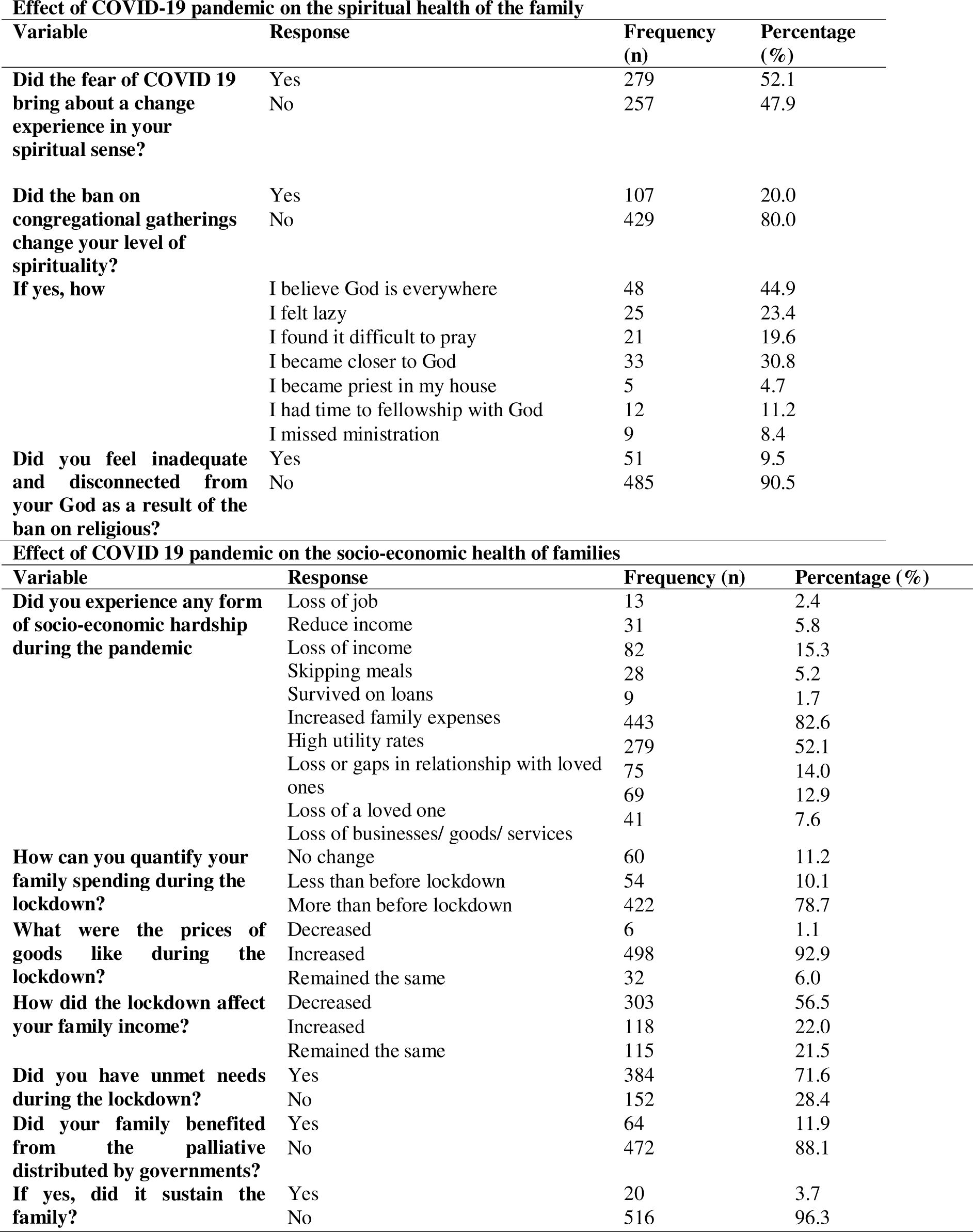
EFFECT OF COVID 19 PANDEMIC ON THE SPIRITUAL HEALTH AND THE SOCIO-ECONOMIC OF THE FAMILY.

## Discussions

The aim of this study is to determine the effect of the COVID-19 and the lockdown on the physical, mental, and socioeconomic health of families

### Effect of Coronal virus disease and its associated lockdown on families’ physical health

This study shows that only 17% of the respondents claimed that COVID-19 affected their family physically, this finding is invariant with a study carried out by Dong et al (2020) which found that health and illness such as Covid-19 is a “family affair”, and clients, Patients, residents and other family members are inextricably connected in the management of the disease. Although not all are infected with COVID-19, but enormous population of individuals and families are affected. This result may be largely due to poor testing and low proportion of people who tested for COVID-19. Also, Dong et al 2020 found that the coronavirus pandemic is causing health problems, discomfort, and pain among families around the world (Dong et al 2020).

With almost 108,399,915 million confirmed cases, over 2.381,134 million deaths, and spreading in 219 countries, no doubt, COVID-19 has shown a tremendous effect on the physical health of individuals and families (WHO, 2021). To begin, the present study shows that only a few of the participants claimed their family members screened for COVID-19, while only 1.5% was tested positive for COVID-19. This finding depicts the low-level of testing among respondents and their families. Furthermore, a modest numbers of the family had a pre-existing medical condition, but 13.6% of the respondents admitted or treated for another form of illness during the pandemic.

Malaria was the highest 61.4% of the illnesses a family member admitted or treated for, equally, 91.4% experienced headache among the family admitted or treated, and only 3.4% of the respondents agreed that COVID-19 pandemic worsened there medical condition. However in a study by Masataka et al. 2020 which opined that, in terms of physical effect on human systems, already known cardiovascular disease patient is associated with various degrees of complications and increase risk of death in patients with the diseases, but coronavirus itself can also induce acute coronary syndrome, myocardial injury, venous thromboembolism, and arrhythmia.

### Effect of coronavirus disease (Covid-19) on Families’ Emotional Health

This present study reveals that families experience anxiety, depression, sleep problem, and fear during the COVID-19 pandemic and it is associated period of lockdown. This was supported by the preliminary evidence found by Rajkumar (2020) found that during the COVI-19 lockdown people presented with the sign and symptoms such as depression, anxiety, self-reported stress, and disturbed sleep, and so on, which are common emotional and psychological reactions to unpleasant events. A number of individual and structural variables moderate this risk. Meanwhile, in another study, Kontoangelor et al., (2020) study on the emotional health effects of Covid-19 pandemic found that children experience anxiety or worry, and fear during COVID-19 pandemic. They also discovered that older adult with underlying health challenges are more susceptible to COVID-19, and this is supported by a study that found that 50.2% of adults experienced anxiety and depression although children were not part of the respondents in the study.

This study also found that during the pandemic, the most family claimed that the lockdown improved their family bonding; this may be corroborated with Rubin and Wessely (2020) study that observed that during the pandemic, families anxiety increases as a result of the following reasons: increase death rate, frighten media broadcast, and an increasing number of confirming new cases. Thus, the stay-at-home order raises anxiety levels. Also, Barbisch et al. (2015) describe how the confinement or stay at home orders result into hysteria condition among families, and this explained the reason for a large number of respondents (74.6%) that claimed improved family bonding.

### Impact of Coronavirus on the families’ spiritual health

COVID-19 pandemic no doubt has taken its toll, either negatively or positively effect on the spirituality of individuals, family and the Nations. Some people have taken their refuge in God and drawn closer to Him and vice versa. This study revealed that a little more than half of the respondents, 52.1% expressed that fear of COVID-19 bring about a change experience in their spiritual sense and them developed personal relationship with their God, this is further explained by the fact that 4.7% of the participants have become priests in their homes, this claim is in agreement with the findings of Egunjobi (2020), who noted that the effect of COVID-19 touches directly on the caring duties, responsibilities, as well as the livelihood of pastors, imams, gurus, rabbis, and other religious leaders to move closer to God. Our study also revealed that few of the respondents, 30.8% claimed becoming closer to God due to the pandemic and lockdown, this result corresponds with a study that found that the pandemic promote religion and trust in God (Pirutinsky et al., 2020).

This study also reveal that, the pandemic had a negative effect on 79.9% of the family’s spiritual health in that, the ban on spiritual gathering resulted in a feeling of inadequacy and disconnectedness from God. It also heightened the fear of the pandemic as equally reported in a study by Moore et al., (2020).

### Impact of Covid-19 on socio-economic health of families

The COVID-19 pandemic continues to affect the socioeconomic health of the family in an unprecedented way. Most of the families spent more than usual during the lockdown, our study corroborated this with 82.6% that they experienced an increase in family expenses, similar studies also reported same in the southwest (Lagos state) and southeast (Anambra and Enugu states) Nigeria (Odii et al., 2020). In addition, there was an increase in the process of goods and foods needed for sustenance, our study revealed that 92.9% of the families reported an increase in goods prices, same was also reported in Ghana (Asante & Helbrecht, 2020), this could be attributed to the passive business operation and possible fear and panic from the business owners (Ozili, 2020).

This study found that during the pandemic just 2.4% of the respondents reported having lost their job, this seems to be very few in relation to a study carried out in Nigeria (Oseni et al., 2020), although there was an increase unemployment level in United State during the lockdown (Bick & Blandin, 2020) this disparity could be attributed to the fact that this study was done a post lockdown period and many respondents reinstated in their places of work. Our study revealed that COVID 19 pandemic had unfavorably effect on 77.2% of the family’s socio-economic health status which consists of the financial capacity, feeding status, expenses, and socialization. To relieve the families of the increase in feeding expenses the Government provided palliative (Eranga, 2020; Ezeah, 2020), our study revealed low beneficiaries of the palliative (11.9%), this was also corroborated with findings which state that the sharing of the palliatives were not evenly distributed (Awofeso & Irabor, 2020; Ezeah, 2020)

This study was intended to assess the effect of COVID-19 on the health of families living in the South-West, Nigeria. The aims of the study were to assess the effect of COVID-19 on the physical health, emotional health, spiritual health, and socioeconomic health of families living in the South-West, Nigeria. To achieve this, four research questions were raised. A descriptive design was used in this study. The subjects of the study comprise (536) five hundred and thirty six participants. The instrument for data collection was self-structured questionnaires, and the questionnaire was divided into five sections (A, B, C, D & E). It consists of thirty-seven questions in all. To ensure the reliability of the questionnaire, a pilot study of the instrument was done through the split-half method and Cronbach’s Alpha was used to test the internal consistency of the instrument. The Cronbach Alpha value was 0.8 was obtained. The final instruments were administered through the use of Google forms. The findings showed that COVID 19 Pandemic had a major effect on the Physical Health of the family, as the study showed that, at this time most of the family members were admitted for Malaria and also noted that COVID 19 pandemic worsened their medical condition. It was also found that the lockdown improved their family bonding, which suggested that COVID 19 pandemic favorably affects the respondents’ emotional health. Moreover, it reveals that COVID 19 pandemic negatively affects the respondents’ spiritual health,. this they said because the ban on congregational gatherings changes their level of spirituality. Furthermore, this study also showed that the COVID-19 pandemic had a negative effect on the socio-economic health of the family. The study shows that there is increased in family expenses such as an increase in utility consumption, that is family’s spending during the lockdown was more than before lockdown and also show that the prices of goods increased during the lockdown.

### Implication for nursing and health policy

Nurses working in COVID-19 unit have an obligation to give total care to the affected patient; therefore, they need to include physical, emotional, spiritual and mental intervention in their care. The government have got to plan and strategies properly in the method for distribution of the palliative’s, and identify the vulnerable and less privileged in each state for easy distribution.

## Conclusion

The present study examined the effect of the COVID-19 and the lockdown on the physical, emotional, spiritual, and socio-economic health of families. It was found that the pandemic has a great effect on the health of families. Therefore the authors recommend the following in view of future occurrences; the government has an obligation to establish workable systems that can quickly respond to pandemics in the future. Spiritually, each family needs to build up their family members to be able to sustain with or without physical gathering and Families need to make up time for regular bonding has experienced during the lockdown. As bonding helps to nurture the family and give room for good parenting and spousal bonding

## Limitations

In this study, the following limitation applicable; even though, the research aim was achieved. The study was limited to only smart phone subscribers who are available online as at the period of data collection and time was also a major limitation face

## Data Availability

All data produced in the present work are contained in the manuscript

## Acknowledgement

The authors wish to sincerely thank all the lecturers of post graduate school Babcock university. Illisha remo, ogun state and our supervisor prof. M.O Akpa.

